# Used paper tissues for pathogen identification in acute respiratory infection

**DOI:** 10.1101/2023.03.24.23287683

**Authors:** Annabel Rector, Mandy Bloemen, Marc Van Ranst, Elke Wollants

## Abstract

We investigated the potential of used paper tissues as a non-invasive sampling method for the diagnosis of acute respiratory infections. The method allowed the identification and typing of respiratory pathogens in symptomatic individuals, as well as in collective samples taken at a community level. The collection of used paper tissues could therefore be useful in epidemiological surveillance for SARS-CoV-2 and other respiratory pathogens such as influenzavirus, respiratory syncytial virus, entero/rhinoviruses and *Streptococcus pneumoniae*.

---

Acute respiratory tract infections (ARIs), including pneumonia, constitute a major disease burden worldwide, especially in young children and the elderly [1][2]. Diagnostic testing for respiratory pathogens is usually performed on samples collected by invasive methods, such as nasopharyngeal swabs, nasopharyngeal aspirates or bronchoalveolar lavages, obtained in hospital or medical practice settings. For some respiratory viruses such as severe acute respiratory syndrome coronavirus 2 (SARS-CoV-2), influenza virus and respiratory syncytial virus (RSV), fast diagnosis using antigen rapid diagnostic test (Ag-RDTs) can be performed on self-collected nasal swabs. Since some of the most vulnerable populations for ARI outbreaks, such as residents of long-term care facilities for the elderly or mentally impaired, or infants and toddlers attending day-nurseries, are difficult to sample using these invasive methods, there is a need for less to non-invasive methods for respiratory sampling. We assessed whether paper tissues, used for nose blowing, can be used for the identification of respiratory pathogens, on an individual as well as on a community level.

## Pathogen identification from used paper tissues

Used paper tissues (UPT) of eight individuals with symptoms of ARI were investigated for the presence of respiratory pathogens. Tissues were transferred with sterile tweezers into a 100 mL disposable syringe (∼4 tissues). Phosphate buffered saline (PBS) was added until tissues were soaked (∼25 mL). After incubation at room temperature for 5 minutes, the plunger of the syringe was pressed to recover the eluate (∼ 10 mL) into a 15 mL Falcon tube. Viral nucleic acids were extracted using the MagMAX™ Viral/Pathogen Nucleic Acid Isolation Kit on Kingfisher Flex-96 (ThermoFisher Scientific, Europe), using 400 μL of eluate of the used paper tissues. Screening for respiratory pathogens was done using an in-house developed respiratory panel (RP) for simultaneous detection of 22 respiratory viruses (influenza A, influenza B, respiratory syncytial virus (RSV), human metapneumovirus, parainfluenzavirus -1 to -4, adenovirus, human bocavirus, enterovirus/rhinovirus (EV/RV), enterovirus D-68, human parechovirus, human coronavirus (HCoV)-NL63, -229E, -OC43, -HKU-1, -SARS and – MERS, cytomegalovirus, herpes simplex virus -1 and -2) and 7 non-viral pathogens (*Mycoplasma pneumoniae, Coxiella burnetii, Chlamydia pneumoniae, Chlamydia psittaci, Streptococcus pneumoniae, Legionella pneumophila* and *Pneumocystis jirovecii*) [3]. An additional SARS-CoV-2 specific RT-qPCR was carried out on samples positive for HCoV-SARS using the 2019-nCoV CDC EUA kit N1 primer probe set as described earlier [4]. Tissues were stored at room temperature for up to 10 days prior to analysis.

In 2 cases, EV/RV was detected, which could be further typed as RV C in one case and as co-infection of RV B and cocksackievirus A19 in the other case (typing performed as described by Wollants et al. [5]). Three samples tested positive for RSV, all three with concomitant detection of an additional pathogen (HCoV-OC43, adenovirus and *Streptococcus pneumoniae* respectively). In one sample, HCoV-OC43 was detected in combination with *Streptococcus pneumoniae*. One sample tested positive for SARS-CoV-2 (Cq 28.1), and was further typed by complete genome Oxford Nanopore Technologies sequencing as variant BA.5.2.1, with a genome coverage of 99.3% [6].

## Respiratory pathogens detected in UPT versus nasal swabs and rapid antigen tests

To further investigate the potential of UPT in comparison to standard diagnostic sampling, UPTs of 16 patients with symptoms of ARI were analysed in parallel with self-collected nasal swabs in Universal Transport Medium (UTM) (Copan). Fourteen of these patients performed an Ag-RDT on the same day. Results are listed in Table 1. In general, pathogens that were detected in the nasal swabs were also detected in the corresponding UPT. Exceptions were HSV-1 (Cq 31.9 in nasal swab) in a patient who was also influenza A positive in both nasal swab and UPT (RP008), and EV/RV (Cq 36.6 in nasal swab) in a patient who also tested positive for influenza B in nasal swab and UPT (RP015). The Cq measured in the UPT was usually higher than in the nasal swab, although in some cases it was the other way round. In all cases where the Ag-RDT tested positive, the corresponding pathogen was also detectable in the UPT.

**Table 1.**
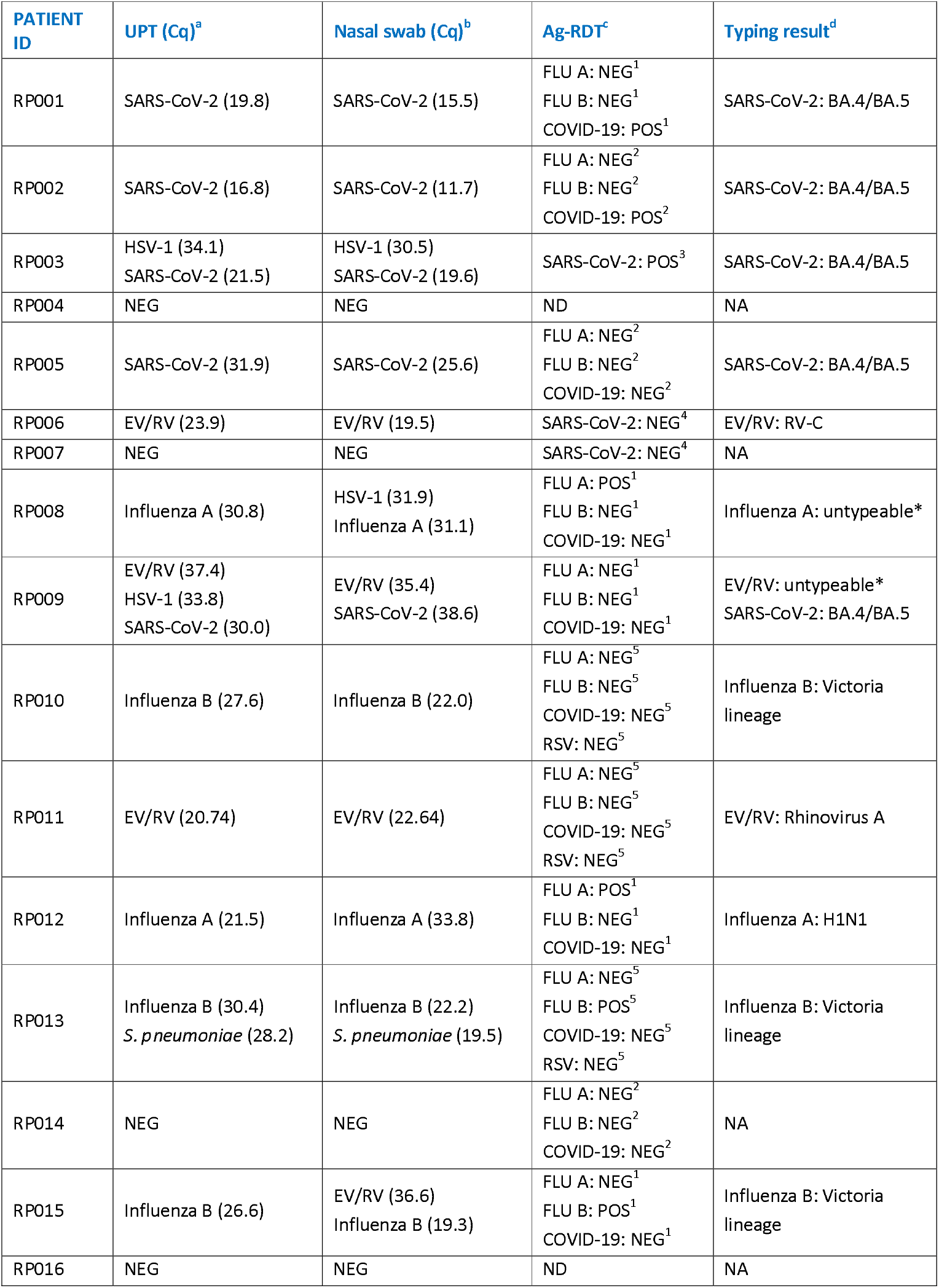
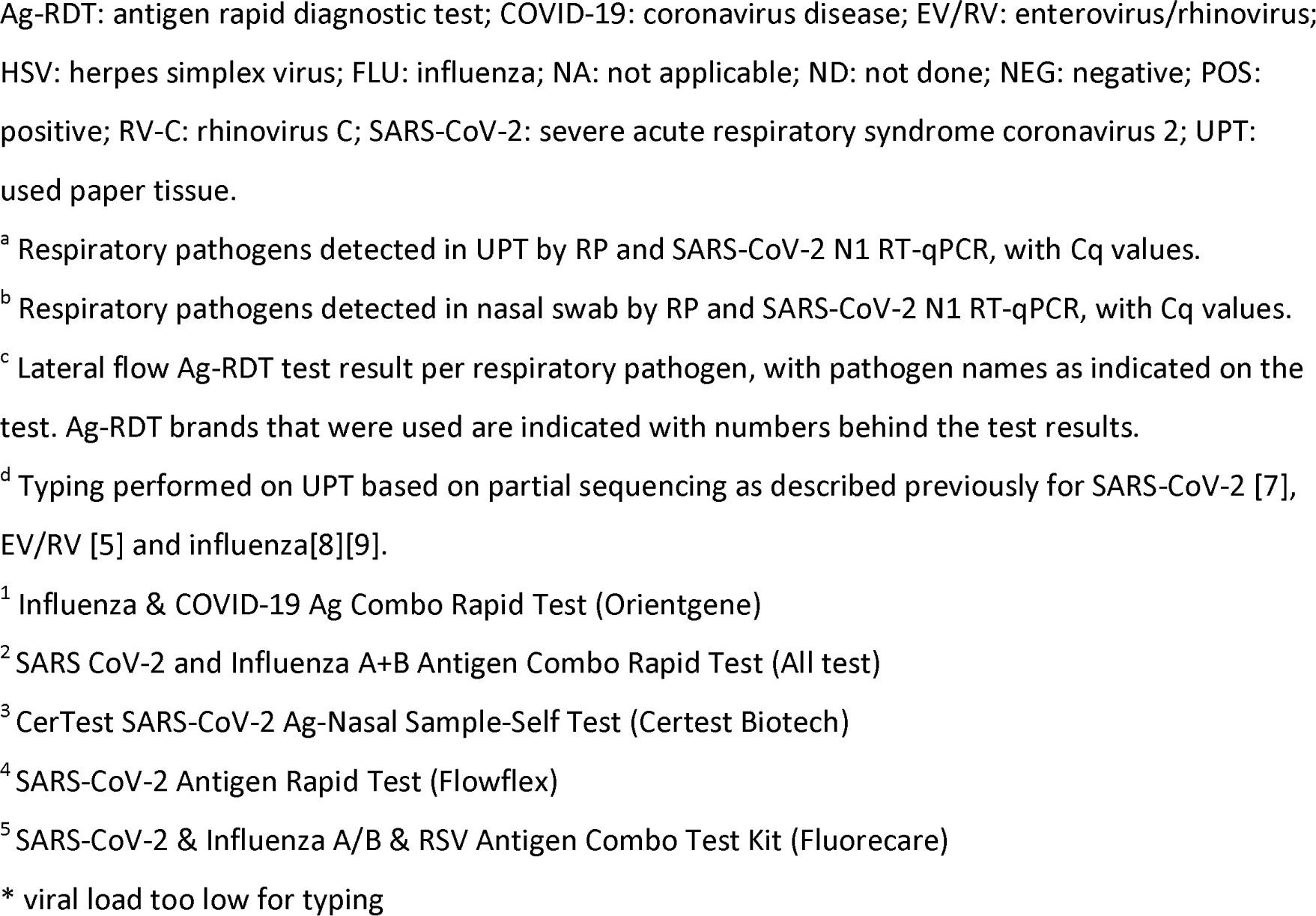
Identification of respiratory pathogens in used paper tissues versus nasal swabs and antigen tests, Belgium, winter 2022-2023 (n□=□16 samples)

| PATIENT ID | UPT (Cq)<sup>a</sup> | Nasal swab (Cq)<sup>b</sup> | Ag-RDT<sup>c</sup> | Typing result<sup>d</sup> |
| --- | --- | --- | --- | --- |
| RP001 | SARS-CoV-2 (19.8) | SARS-CoV-2 (15.5) | FLU A: NEG<sup>1</sup><br>FLU B: NEG<sup>1</sup><br>COVID-19: POS<sup>1</sup> | SARS-CoV-2: BA.4/BA.5 |
| RP002 | SARS-CoV-2 (16.8) | SARS-CoV-2 (11.7) | FLU A: NEG<sup>2</sup><br>FLU B: NEG<sup>2</sup><br>COVID-19: POS<sup>2</sup> | SARS-CoV-2: BA.4/BA.5 |
| RP003 | HSV-1 (34.1)<br>SARS-CoV-2 (21.5) | HSV-1 (30.5)<br>SARS-CoV-2 (19.6) | SARS-CoV-2: POS<sup>3</sup> | SARS-CoV-2: BA.4/BA.5 |
| RP004 | NEG | NEG | ND | NA |
| RP005 | SARS-CoV-2 (31.9) | SARS-CoV-2 (25.6) | FLU A: NEG<sup>2</sup><br>FLU B: NEG<sup>2</sup><br>COVID-19: NEG<sup>2</sup> | SARS-CoV-2: BA.4/BA.5 |
| RP006 | EV/RV (23.9) | EV/RV (19.5) | SARS-CoV-2: NEG<sup>4</sup> | EV/RV: RV-C |
| RP007 | NEG | NEG | SARS-CoV-2: NEG<sup>4</sup> | NA |
| RP008 | Influenza A (30.8) | HSV-1 (31.9)<br>Influenza A (31.1) | FLU A: POS<sup>1</sup><br>FLU B: NEG<sup>1</sup><br>COVID-19: NEG<sup>1</sup> | Influenza A: untypeable* |
| RP009 | EV/RV (37.4)<br>HSV-1 (33.8)<br>SARS-CoV-2 (30.0) | EV/RV (35.4)<br>SARS-CoV-2 (38.6) | FLU A: NEG<sup>1</sup><br>FLU B: NEG<sup>1</sup><br>COVID-19: NEG<sup>1</sup> | EV/RV: untypeable*<br>SARS-CoV-2: BA.4/BA.5 |
| RP010 | Influenza B (27.6) | Influenza B (22.0) | FLU A: NEG<sup>5</sup><br>FLU B: NEG<sup>5</sup><br>COVID-19: NEG<sup>5</sup><br>RSV: NEG<sup>5</sup> | Influenza B: Victoria lineage |
| RP011 | EV/RV (20.74) | EV/RV (22.64) | FLU A: NEG<sup>5</sup><br>FLU B: NEG<sup>5</sup><br>COVID-19: NEG<sup>5</sup><br>RSV: NEG<sup>5</sup> | EV/RV: Rhinovirus A |
| RP012 | Influenza A (21.5) | Influenza A (33.8) | FLU A: POS<sup>1</sup><br>FLU B: NEG<sup>1</sup><br>COVID-19: NEG<sup>1</sup> | Influenza A: H1N1 |
| RP013 | Influenza B (30.4)<br>*S. pneumoniae* (28.2) | Influenza B (22.2)<br>*S. pneumoniae* (19.5) | FLU A: NEG<sup>5</sup><br>FLU B: POS<sup>5</sup><br>COVID-19: NEG<sup>5</sup><br>RSV: NEG<sup>5</sup> | Influenza B: Victoria lineage |
| RP014 | NEG | NEG | FLU A: NEG<sup>2</sup><br>FLU B: NEG<sup>2</sup><br>COVID-19: NEG<sup>2</sup> | NA |
| RP015 | Influenza B (26.6) | EV/RV (36.6)<br>Influenza B (19.3) | FLU A: NEG<sup>1</sup><br>FLU B: POS<sup>1</sup><br>COVID-19: NEG<sup>1</sup> | Influenza B: Victoria lineage |
| RP016 | NEG | NEG | ND | NA |

Ag-RDT: antigen rapid diagnostic test; COVID-19: coronavirus disease; EV/RV: enterovirus/rhinovirus; HSV: herpes simplex virus; FLU: influenza; NA: not applicable; ND: not done; NEG: negative; POS: positive; RV-C: rhinovirus C; SARS-CoV-2: severe acute respiratory syndrome coronavirus 2; UPT: used paper tissue.
<sup>a</sup> Respiratory pathogens detected in UPT by RP and SARS-CoV-2 N1 RT-qPCR, with Cq values.
<sup>b</sup> Respiratory pathogens detected in nasal swab by RP and SARS-CoV-2 N1 RT-qPCR, with Cq values.
<sup>c</sup> Lateral flow Ag-RDT test result per respiratory pathogen, with pathogen names as indicated on the test. Ag-RDT brands that were used are indicated with numbers behind the test results.
<sup>d</sup> Typing performed on UPT based on partial sequencing as described previously for SARS-CoV-2 [7], EV/RV [5] and influenza[8][9].
<sup>1</sup> Influenza & COVID-19 Ag Combo Rapid Test (Orientgene)
<sup>2</sup> SARS CoV-2 and Influenza A+B Antigen Combo Rapid Test (All test)
<sup>3</sup> CerTest SARS-CoV-2 Ag-Nasal Sample-Self Test (Certest Biotech)
<sup>4</sup> SARS-CoV-2 Antigen Rapid Test (Flowflex)

## SARS-CoV-2 load in UPT versus nasal swab and Ag-RDT over the course of infection

One patient was followed over the course of COVID infection, from the time of first symptom up to complete symptom resolution, with UPTs, nasal swabs in UTM and Ag-RDTs being collected daily (Fig). SARS-CoV-2 was detectable in UPT as of the start of symptoms, whereas the Ag-RDT only turned positive on Day 4. For as long as the Ag-RDT remained positive, SARS-CoV-2 was also detectable in UPT. In the nasal swab, SARS-CoV-2 was still detectable after Ag-RDTs turned negative, and remained detectable up to the last symptomatic day.

**Figure.**
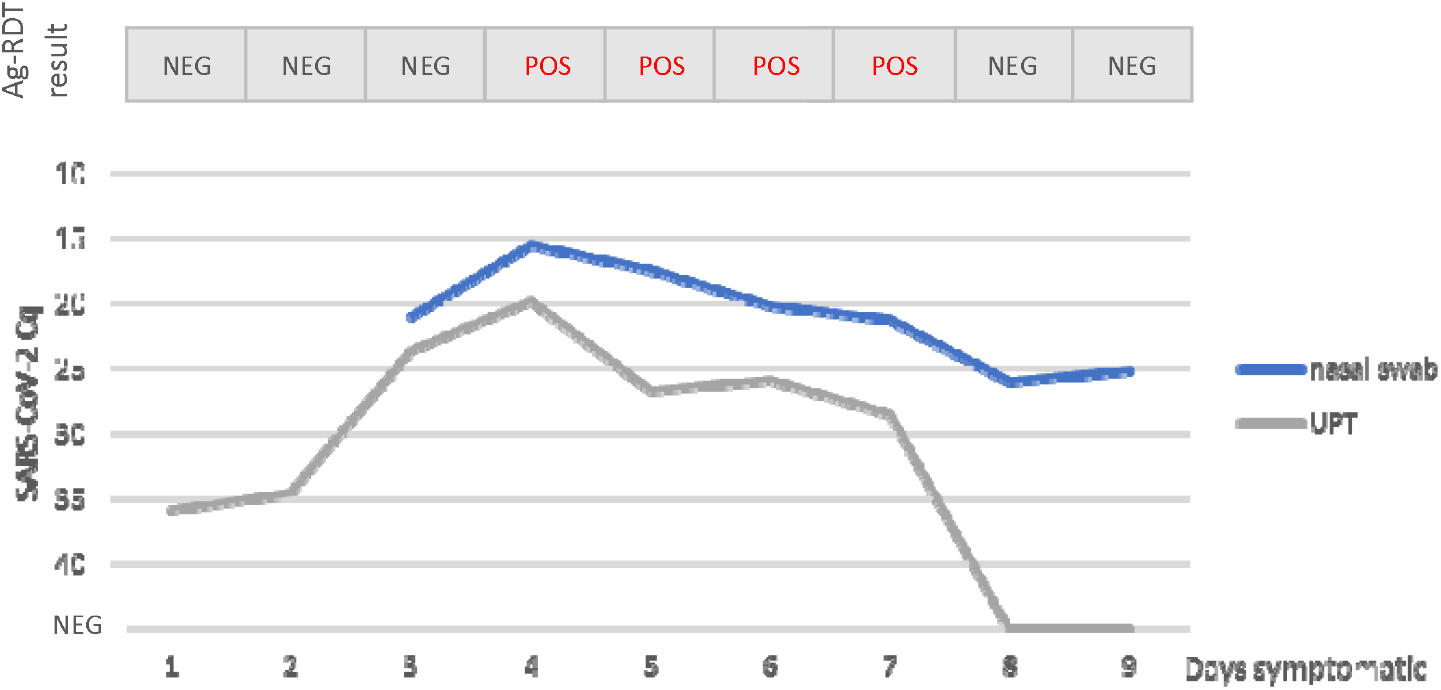
Evolution of SARS-CoV-2 load over the course of an infection, as captured by Cq values in UPT and nasal swab, and Ag-RDT test result, Belgium, February 2023.

## Respiratory pathogens in combined UPTs of collectivities

Combined UPTs from 6 collectivities (one childcare centre, 3 kindergartens and 2 primary schools) were collected by anonymously gathering tissues used in a classroom or childcare group over the course of one day. The maximal amount of tissues was used for pathogen elution, and further investigated as described above. The presence of multiple respiratory pathogens was detected in these combined samples (Table 2).

**Table 2.**
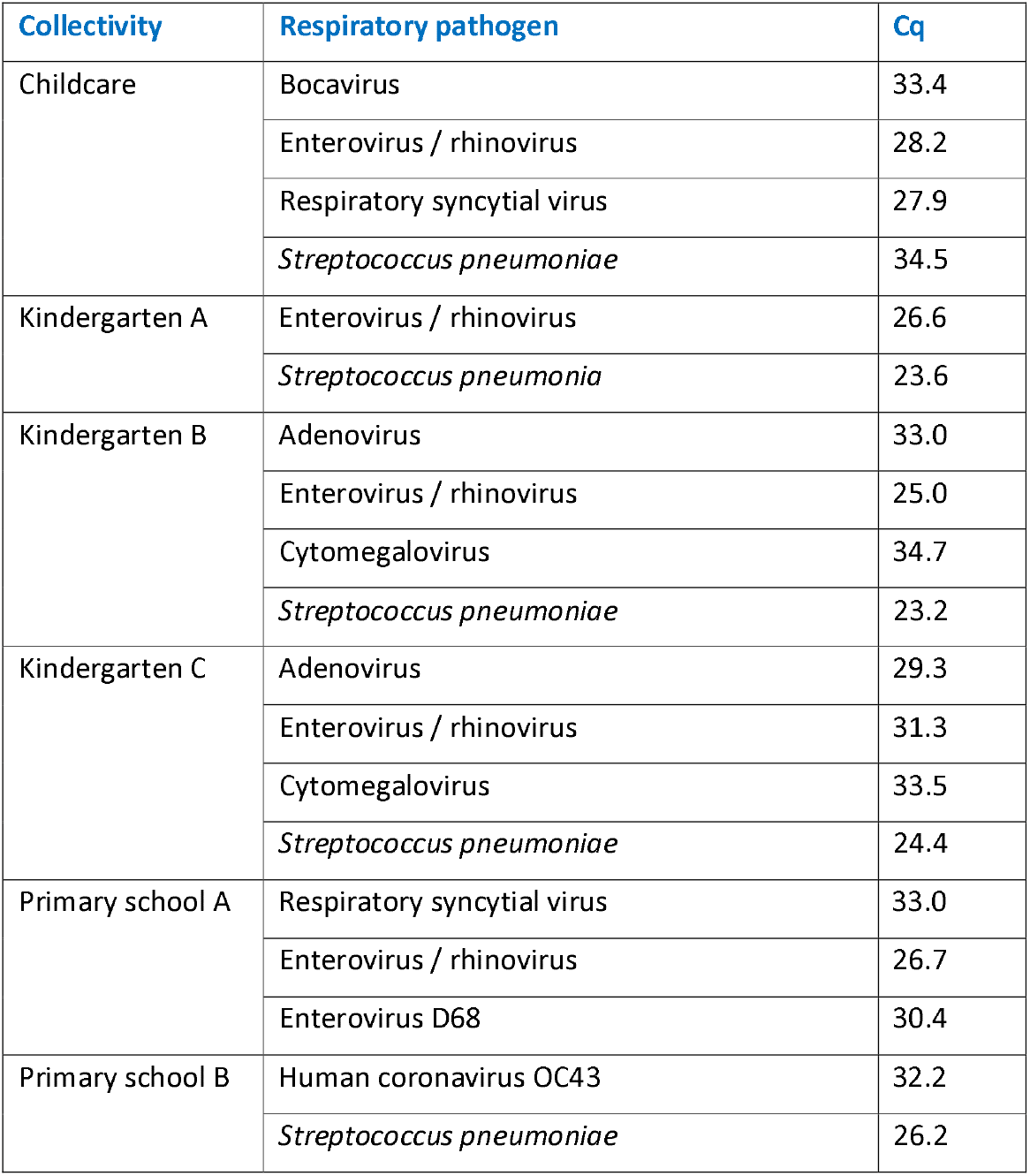
Identification of respiratory pathogens in combined used paper tissues from collectivities, Belgium, November 2022 (n□=□6 samples)

## Discussion

Our method for detection of respiratory pathogens from UPT samples allowed the identification of respiratory pathogens responsible for ARI, with adequate sample quality to allow further genetic characterization. We were able to detect respiratory viruses as well as bacteria, with most pathogens that were identified in nasal swabs also being detected in concurrent UPTs from the same patient. Pathogens that were not detected in the UPT samples were co-infecting pathogens which were only present in low amounts in the corresponding nasal swab (Cq ≥ 32).

Bacterial pathogens have been shown to be reliably detectable from paper tissues of patients with upper respiratory tract infections [10]. In a recent study, Lagathu et al. were able to identify multiple respiratory viruses in pooled facial tissues obtained in communities of children. They also compared SARS-CoV-2 Cq values between nasopharyngeal swabs and facial tissues of individual COVID-19 patients and found a higher signal from the tissues in 11 out of in 15 cases [11]. In our study, we compared Cq values for SARS-CoV-2 but also for other common respiratory viruses such as enterovirus/rhinovirus and influenzavirus A and B, and *S. pneumoniae*, obtained from 16 UPT and nasal swabs, and found a high variety in Cq difference between both samples. We also were able to detect the presence of multiple respiratory pathogens in pooled UPT samples of collectivities, confirming its applicability for community testing. This would especially be useful in schools and preschool daycare centers, since taking nasal samples from (young) children is an invasive method and requires training, or in elderly homes and homes for disabled people, in whom taking nasal samples is less well tolerated. Because sequencing a complete genome is possible from UPT this method can also be applied for epidemiological surveillance. Furthermore, UPT samples can easily be transported to diagnostic laboratories, even by regular mail.

Since our sample contains eluted material from entire paper tissues, the pathogen load in the sample is not only dependent on the amount of virus shedding but also on the amount of nasal discharge collected in the tissue. This makes the method less suited for (semi-) quantitative analyses. It also implies that the method cannot be used when there is very little to no nasal discharge, or when nasal discharge is difficult to collect by nose blowing or wiping with a tissue.

We were able to detect the corresponding virus in UPT of all Ag-RDT positive cases, indicating that UPTs are sufficiently sensitive to detect individuals with high virus shedding, who are most likely to be infectious. As such, UPT could provide an interesting non-invasive sampling method for screening of individuals. In the patient that was followed over the course of a COVID infection, UPTs tested positive as of the start of symptoms, whereas Ag-RDTs turned positive only on day 4. This is in accordance with the notion that SARS-CoV-2 viral loads in persons with pre-existing immunity (by previous infection or by vaccination) may only rise to Ag-RDT detectable levels after several days of symptoms. Although only based on a single observation, UPT testing seems to be sensitive enough to allow detection as of the start of infection, reducing the amount of false negative test results.

Since pathogen detection was possible from combined UPTs obtained in collectivities, it can also provide a good alternative to sampling of sewage water of buildings or aircraft wastewater to obtain a community sample for pathogen screening. This would be very useful to complement the current strategy of wastewater testing of incoming aircraft for SARS-CoV-2 variant screening [12], [13].

## Data Availability

All data produced in the present study are available upon reasonable request to the authors

## Ethical statement

Ethics committee of UZ/KU Leuven gave ethical approval for this work.

Informed consent, approved by the UZ Leuven Ethics Committee, was obtained from all individuals providing self-collected samples.

## Acknowledgements

UZ Leuven, as national reference center, is supported by Sciensano.

## Conflict of interest

None declared.

## Authors’ contributions

Annabel Rector: Conception and design of the study, data analysis and interpretation, writing of the manuscript, final approval of the version to be published.

Mandy Bloemen: Conception and design of the study, data collection, data analysis and interpretation, writing of the manuscript, final approval of the version to be published.

Marc Van Ranst: Conception and design of the study, final approval of the version to be published.

Elke Wollants: Conception and design of the study, data collection, data analysis and interpretation, writing of the manuscript, final approval of the version to be published.

## Notes

### Competing Interest Statement

The authors have declared no competing interest.

### Funding Statement

This study did not receive any funding

### Author Declarations

Ethics committee of UZ/KU Leuven gave ethical approval for this work (S67382). Informed consent, approved by the UZ Leuven Ethics Committee, was obtained from all individuals.

